# Surgical complications during pregnancy following bariatric surgery: a Belgian nationwide population-based study

**DOI:** 10.64898/2026.03.30.26349694

**Authors:** Paulien De Mulder, Karolien Benoit, Caroline Daelemans, Fréderic Debiève, Roland Devlieger, Kristien Roelens, Yves Van Nieuwenhove, Griet Vandenberghe, the B.OSS Collaborating Group

## Abstract

**Objective:** To determine the incidence and clinical characteristics of surgical complications during pregnancy in women with a history of bariatric surgery.

**Design:** A nationwide, prospective, population-based cohort study.

**Setting:** High-risk obstetric care in Belgium: 67.6% of maternity units participated, covering 65% of all births in the study period.

**Participants:** Pregnant women with a history of bariatric surgery presenting with a surgical complication (internal hernia, intussusception, volvulus or adhesions; anastomotic ulcer or abscess; gastric band slippage; or incisional hernia) between January 2021 and December 2022.

**Results:** Thirty-three women experienced 35 surgical complications. Internal herniation was most common (n=25), predominantly following Roux-en-Y gastric bypass. Mean gestational age at diagnosis was 27+6 weeks. All women underwent surgical exploration within 24 hours; bowel resection was required in two cases. Caesarean section occurred in 48.5%, with 13 preterm births and one neonatal death. One woman required intensive care. No maternal death occurred.

**Conclusion:** Surgical complications following bariatric surgery in pregnancy are uncommon but carry significant obstetric risks. All observed complications occurred after procedures involving intestinal rerouting, predominantly Roux-en-Y gastric bypass. Prompt surgical management was associated with low maternal morbidity and no mortality, but frequently resulted in preterm birth and emergency caesarean section. These findings highlight the need for a low threshold for surgical evaluation of abdominal pain in pregnant women with previous bariatric surgery and suggest that procedure type is relevant when counselling women of reproductive age.

**Strengths and limitations:** *Strengths:* - Nationwide, population-based cohort study to evaluate surgical complications during pregnancy following bariatric surgery. - Case identification was performed through the Belgian Obstetric Surveillance System (B.OSS), using active prospective monthly reporting. Cases were compared with national hospital discharge data to assess potential underreporting. - National health insurance data were used to estimate the total number of pregnancies following bariatric surgery to provide procedure-specific denominators.

*Limitations:* - The small number of cases limited statistical power and precluded formal comparisons between different bariatric procedures. - Despite its strong network, the B.OSS methodology is vulnerable to underreporting due to the voluntary participation of clinicians.

## Introduction

Bariatric surgery (BS) has gained popularity in response to the escalating global epidemic of obesity, defined as a body mass index (BMI) ≥ 30 kg/m². In Belgium, a total of 13 779 bariatric procedures were performed in 2021.^1^ The most established procedures include gastric bypass procedures (72%), sleeve gastrectomy (SG) (27%) and laparoscopic adjustable gastric banding (LAGB) (1%).^2^

Bariatric patients are predominantly female (73%), many of whom are of reproductive age (15-45 years).^2, 3^ This has led to an increase in pregnancies following BS. While BS prior to pregnancy is associated with significant reductions in obesity-related comorbidities, such as preeclampsia and gestational diabetes^3^, it also comes with new challenges due to the anatomical and physiological changes that occur. The altered absorption sites for essential nutrients (iron, calcium, vitamin D, vitamin B12, vitamin A, and folate) may lead to maternal deficits and can affect fetal development.^4, 5^ Furthermore, BS has been associated with an increased risk of complications such as preterm birth, small for gestational age (SGA) infants, congenital anomalies, perinatal mortality, and higher rates of neonatal intensive care unit (NICU) admissions compared to women without prior BS.^6^

Late-onset surgical complications following BS are a recognized clinical concern. When occurring during pregnancy, these complications pose risks to both maternal and fetal health and may be challenging to diagnose, as symptoms often overlap with common pregnancy-related complaints, potentially leading to delayed recognition and treatment.

A systematic review showed evidence for two late-term surgical complications during pregnancy: gastric band slippage following LAGB, and bowel strangulation, mainly following Roux-en-Y gastric bypass (RYGB).^6^ Small bowel strangulation is a recognized complication of BS, especially caused by an internal hernia (IH) after RYGB or one anastomosis gastric bypass (OAGB). The lifetime incidence of IH post-RYGB is up to 10%.^7^ During pregnancy, the incidence is reported at 8%.^8^ The high occurrence is attributed to the physiological enlargement of the uterus (particularly in the third trimester) resulting in increased abdominal pressure and displacement of the intestines.^9^ Other causes of obstruction include intussusception, volvulus, post-operative adhesions or anastomotic strictures. Rapid weight loss that is more than expected is a risk factor for internal herniation, as it may lead to the appearance or reappearance of mesenteric defects.^4, 10^

The incidence of band slippage after LAGB is estimated to be higher (12%) during pregnancy and the postpartum period, compared to the incidence of 3-5% in the general population.^11^ This may be attributed to the increased abdominal pressure associated with repeated vomiting during pregnancy. Band slippage can be resolved with band deflation and repositioning, or surgical removal if gastric strangulation is suspected.^9^ Pregnancy is associated with higher rates of band deflation, and surgical revisions occur more frequently in the postpartum period.^6^

We aimed to assess the incidence of surgical complications after BS during pregnancy and to analyze their clinical manifestations, diagnostic approaches, management strategies, and outcomes for the mother and neonate in a nationwide population-based study.

## Materials and Methods

### Research approach

This study was conducted within the network of the Belgian Obstetric Surveillance System (B.OSS). B.OSS was initiated in 2011 as a national reporting system that investigates severe maternal morbidity in Belgium.^12^ B.OSS is supported by the Federal Public Service of Health and is part of the International Network of Obstetric Survey Systems (INOSS).

### Data collection

A nationwide, prospective population-based surveillance study was conducted using the B.OSS platform. Participation of 67.6% of Belgian maternity units (N = 102) covered 65% (N = 149432) of all births. Cases were collected through monthly active reporting using the ‘nothing-to-report’ principle, with online data collection forms completed for reported cases. The questionnaire was based on the United Kingdom Obstetric Surveillance System (UKOSS) version, allowing international comparisons.^13^ Reminders were sent in case of missing data or incomplete data, up to 12 months after the study period.

To explore potential underreporting, we compared our data against the Belgian hospital discharge register (Minimale Ziekenhuis Gegevens (MZG), Résumé Hospitalier Minimum (RHM)) by combining relevant ICD-10-CM codes: - O9984* (BS status complicating pregnancy) and K95* (complications of bariatric procedures).

The denominator (pregnancies after BS) was obtained from the InterMutualistic Agency (IMA-AIM), which records reimbursed healthcare data of Belgian inhabitants affiliated to the seven Belgian Health Insurance funds. The IMA-AIM database includes information of every medical act that was reimbursed in Belgium based on a specific nomenclature code, such as the timing and location of the medical act and (coded) patient data.(see attachments).

### Ethical approval

The research proposal was approved by the Central Ethics Committee at Ghent University Hospital (EC number B670201526875). Local ethic committee approvals were obtained. Patients provide informed consent and could opt-out at any time. Data were anonymised.

### Case definition

Patients eligible for inclusion were defined as pregnant women, identified with BS prior to conception, presenting with a surgical complication defined as internal herniation (with or without obstructive features), intussusception, volvulus, anastomotic ulceration or abscess, gastric band erosion or slippage, and ventral incisional hernia.

### Registered variables

The survey requested information on maternal demographics and medical history, as well as details about the BS, obstetric history, current pregnancy and circumstances of the surgical complication. It also asked about the management of the complication, and the maternal and neonatal outcomes. Surgical reports were requested, to obtain more anatomical details about the complication.

### Study period

We included cases that occurred between January 2021 and December 2022.

### Statistical analysis

We estimated the incidence of BS-related surgical complications during pregnancy by taking the total number of pregnant women with a history of BS during the study period retrieved from IMA-AIM database as the denominator. A full description of the necessary data manipulations and statistical analysis is included in the supplementary material. Odds ratios (OR) and 95% confidence intervals (CI) were calculated in univariate analysis to compare patient and management characteristics. Logistic regression analysis was used to explore associations between the independent variables, treatment plans (e.g. laparoscopy or laparotomy) and obstetric outcomes. Non-parametric tests were applied to compare distributions between groups: the chi-squared test was used for categorical variables and the Mann-Whitney U test for continuous variables. P-values of less than 0.05 were considered statistically significant. Data were analyzed using the IBM SPSS Statistics V.29 statistical software package.

### STROBE cohort guidelines

We used the STROBE writing guide when drafting this article, and the STROBE checklist (see supplementary materials Table 2) to demonstrate adherence to the STROBE reporting guideline.^14^

## Results

Between January 2021 and December 2022, 33 women were included in the study. A further seven women were registered but were excluded from the analysis: two due to missing data in the collection forms, two due to non-surgical complications (pre-eclampsia and serious anemia), and three due to double registration. Two women experienced a second episode of bowel obstruction during pregnancy, resulting in a total of 35 cases of surgical complications being included in the study (Figure 1). Most cases were submitted by hospitals in Flanders (N = 29), followed by three cases from Wallonia and one from the Brussels region.

**Figure 1:**
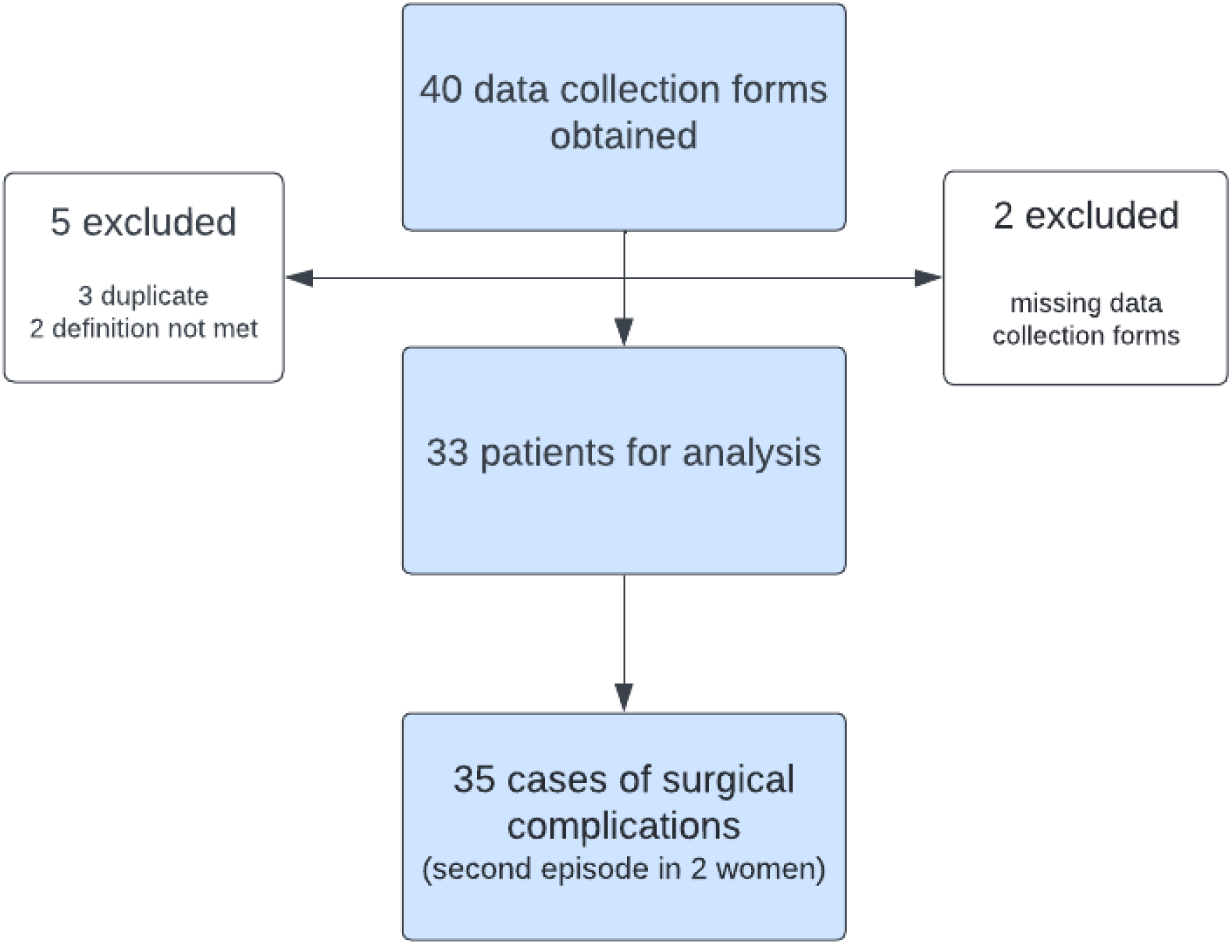
Flowchart of case reporting and data collection.

Using national health insurance claims data (IMA-AIM), we identified a total of 6,217 pregnancies following BS during our study period, including 3,992 pregnancies after RYGB and 2,158 after sleeve gastrectomy. This corresponds to a surgical complication rate of 0.56% after BS in total (95% CI: 0.38-0.75%), while it was higher at 0.88% (95% CI: 0.58-1.16%) in the subgroup of women who had undergone RYGB. No complications were observed after sleeve gastrectomy.

Using the ICD-10-CM codes O9984* (BS status complicating pregnancy) and K95* (complications of bariatric procedures), the MZG-RHM recorded a total of 46 hospital stays for 40 women during the study period, with surgery performed in 91% of cases (N = 42). This indicates a potential underreporting of seven women, and 11 hospital stays within our dataset, though it is important to note that the code K95* also includes non-surgical complications (e.g. dumping syndrome).

### Population description

The mean maternal age at the beginning of the pregnancy was 29.7 years old (95% CI 22.7 to 38.0). The relevant characteristics of the women are displayed in Figure 2. The RYGB was by far the most performed type of BS (N = 29). In two cases, RYGB was performed subsequently after SG due to weight regain after the initial procedure. Regardless of parity, this was the first pregnancy after BS in 20 women. The surgery-to-conception interval was less than one year in five women.

**Figure 2:**
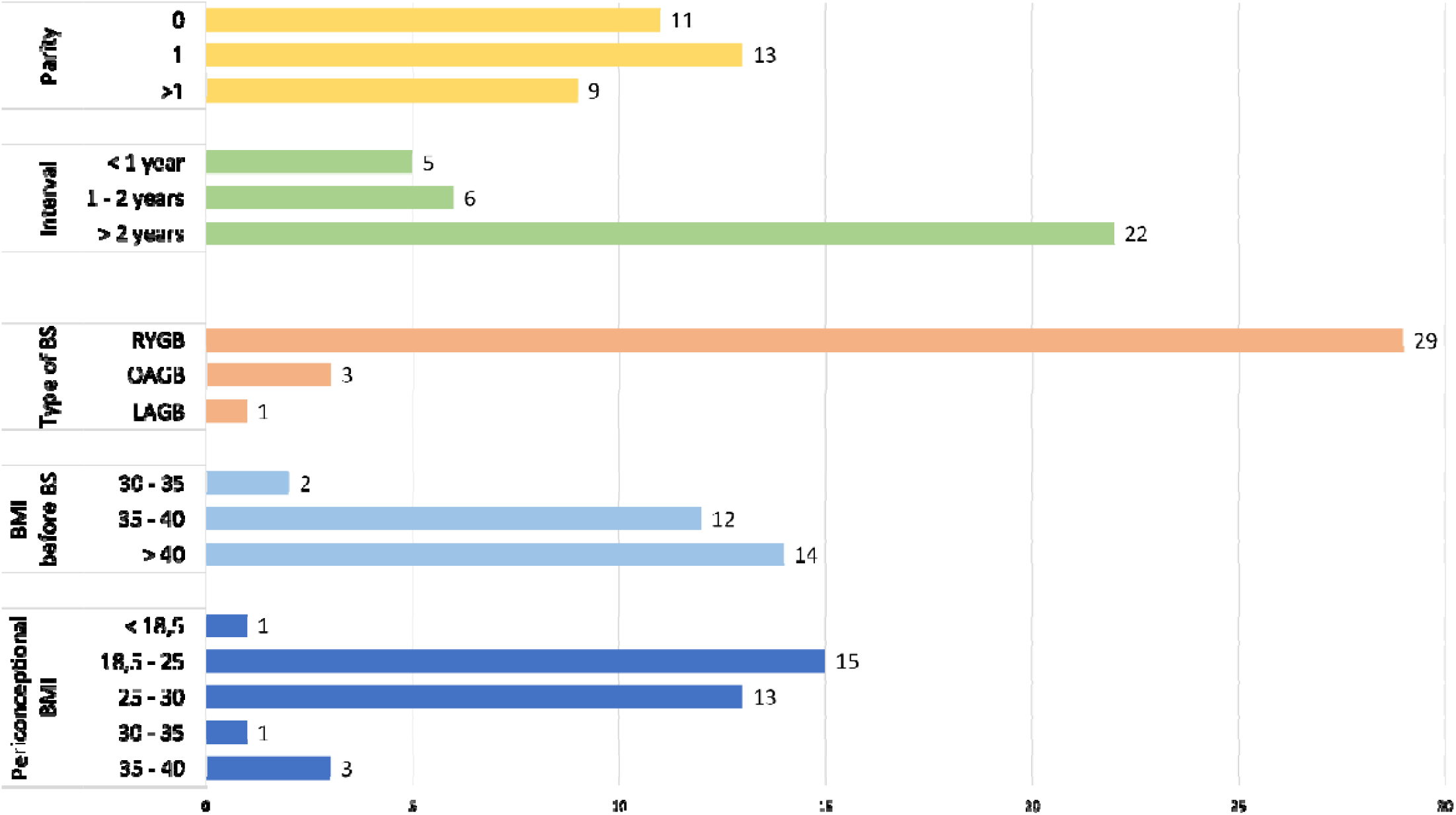
Maternal characteristics. *Abbreviations: BMI = body mass index, BS = bariatric surgery, RYGB = Roux-en-Y-gastric bypass, OAGB = one anastomosis gastric bypass, = laparoscopic adjustable gastric banding. LAGB*

All the women were obese prior to undergoing weight loss surgery (mean BMI 40.6 kg/m2, 95% CI 34.7 to 48.3). The mean total weight loss was 41.3 kg (36.8%; 95% CI 7.6% to 54.2%) at the start of the pregnancy. The mean periconceptional BMI was 25.7 kg/m2 (95%CI 18.4 to 37.3). The medical history of each patient was reviewed. Sixteen women had a history of abdominal surgery, with cholecystectomy being the most frequently performed procedure. Four women who had undergone RYGB experienced an IH before this pregnancy.

### Surgical complications

The mean gestational age at the time of diagnosis of a surgical complication was 27 weeks and 6 days (equivalent to 195 days, 95% CI 87.2 to 266.6). Of these, only one complication occurred during the first trimester, while 15 occurred during the second trimester and 19 occurred during the third trimester.

The details of the surgical complications are shown in Table 1. The most frequent surgical complication was IH with or without obstructive features after bypass procedures (N = 25). In four cases, IH coexisted with either volvulus (N = 2) or intussusception (N = 2).

**Table 1:** Causes of surgical complications. Abbreviations: RYGB = Roux-en-Y-gastric bypass, OAGB = one anastomosis gastric bypass, LAGB = laparoscopic adjustable gastric banding *\** through the mesentery of the Roux limb and the transverse mesocolon (Petersen’s space)

|  |  | **RYGB** | **OAGB** | **LAGB** | **Total** |
| --- | --- | --- | --- | --- | --- |
|  | Petersen* |  |  |  | 9 |
|  | Jejunojejunostomy |  |  |  | 7 |
|  | Not specified |  |  |  | 9 |
| Intussusception |  | 5 | 1 |  | 6 |
| Volvulus |  | 2 | 1 |  | 3 |
| Adhesions |  | 3 |  |  | 3 |
| Gastric band slippage |  |  |  | 1 | 1 |
| Suture abscess |  | 1 |  |  | 1 |
| **Total** |  | **34** | **4** | **1** | **39** |

One non-obstructive emergency after RYGB was noted: a suture abscess at the gastrojejunostomy anastomosis. Notably, the surgery-to-conception interval in this case was almost five years. One patient experienced band slippage after LAGB. Preventive band deflation was not considered during pregnancy.

Nearly all women presented with abdominal pain (N = 34), followed by nausea (N = 23) and vomiting (N = 19). Only four women presented with obstetric symptoms, such as contractions or vaginal bleeding (N = 4). Most women were admitted immediately upon their first hospital presentation (N = 21). Eleven had previously been seen by a healthcare professional but not admitted to hospital: eight by their gynecologist, one by the general practitioner, and two by an emergency physician. Laboratory results at the time of admission were available for 33 cases. Most women (n = 20) had normal blood test results. Considering the pregnancy-adjusted reference intervals ^15–17^, C-reactive protein was elevated (> 20 mg/L) in six women, and four women had leukocytosis (white blood cell count >15 x 10^9^/L). Anemia (Hb <10 g/dL) was observed in a quarter of the women. Lactate levels were measured in 11 cases, with three of them showing elevated levels (>2.2 mmol/L).

Various imaging techniques were used. Table 2 shows the imaging techniques used for all cases with suspected internal hernia or bowel obstruction.

**Table 2:**
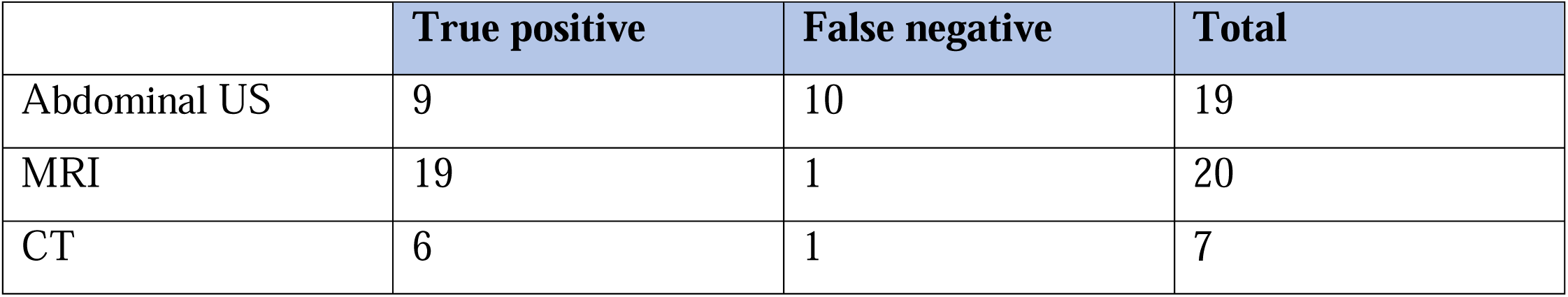
Imaging techniques in women with suspected internal hernia or bowel obstruction.

|  | True positive | False negative | Total |
| --- | --- | --- | --- |
| Abdominal US | 9 | 10 | 19 |
| MRI | 19 | 1 | 20 |
| CT | 6 | 1 | 7 |

The patient with LAGB was assessed with X-ray (with and without barium contrast), CT scan, and diagnostic gastroscopy. The patient with a suture abscess underwent CT scanning with immediate suspicion of an intra-abdominal collection at the gastrojejunostomy.

The management details are presented in Figure 3. Conservative measures consisted of nil per os, analgesia, if indicated, intravenous fluids and antibiotics. Laparoscopic exploration was performed on 21 women, three of whom required conversion to open surgery due to intestinal perforation or unsuccessful laparoscopic reduction of the obstruction. All surgical interventions were performed within 24 hours of admission. Nevertheless, two women with small bowel strangulation and segmental bowel necrosis required resection.

**Figure 3:**
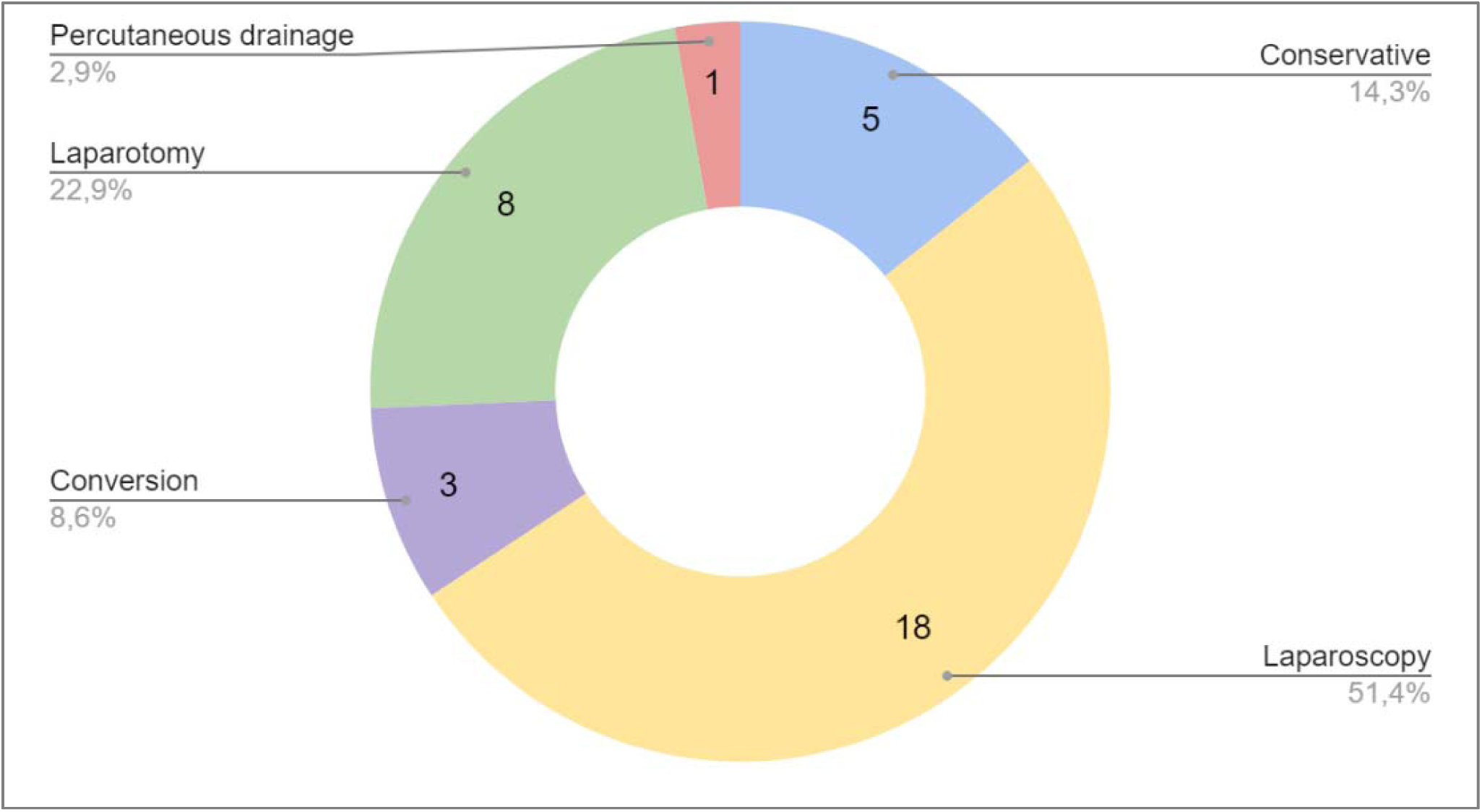
Management of the surgical complication.

One woman with a suture abscess was referred to a tertiary hospital where abscess drainage was performed under image guidance. The case was further complicated by pneumonia with parapneumonic effusion, which necessitated pleural drainage. The woman required admission to the intensive care unit due to sepsis but made a full recovery.

Logistic regression analysis was conducted to assess the influence of independent factors on surgical technique selection. The findings are summarized in Table 3. Variables without a clinically plausible association or meaningful effect estimates were not retained in the final model. Only gestational age showed a marginal association with a greater likelihood of requiring open surgery compared to laparoscopy (OR = 1.02, 95% CI 1.001 to 1.037). In our series, a laparoscopic approach was performed as late as 33 weeks of gestation.

**Table 3:** Logistic regression model for type of surgery.

| Predicting factors for type of surgery | Standard error | Odds ratio | 95% CI |
| --- | --- | --- | --- |
| Gestational age at complication | 0,009 | 1,02 | 1,001 - 1,037 |
| Periconceptional BMI | 0,096 | 0,96 | 0,797 - 1,161 |
| Periconceptional weight | 0,029 | 0,98 | 0,928 - 1,041 |
| History of abdominal surgery | 0,832 | 0,24 | 0,047 - 1,219 |
| Time between symptom onset and admission | 0,018 | 0,98 | 0,948 - 1,019 |

### Obstetric outcomes

The mean gestational age at delivery was 37 weeks and 3 days (range 220 to 284 days). Thirteen women delivered preterm (<37weeks), including one very preterm delivery (<32 weeks). For women who underwent laparotomy, the gestational age at delivery had a broad range (mean 252.2 days; range 220 to 284 days). However, the mean gestational age was lower for women who underwent laparotomy compared to those who underwent laparoscopy (mean 268.7 days; range 245 to 283 days) (P = 0.013) (Figure 4).

**Figure 4:**
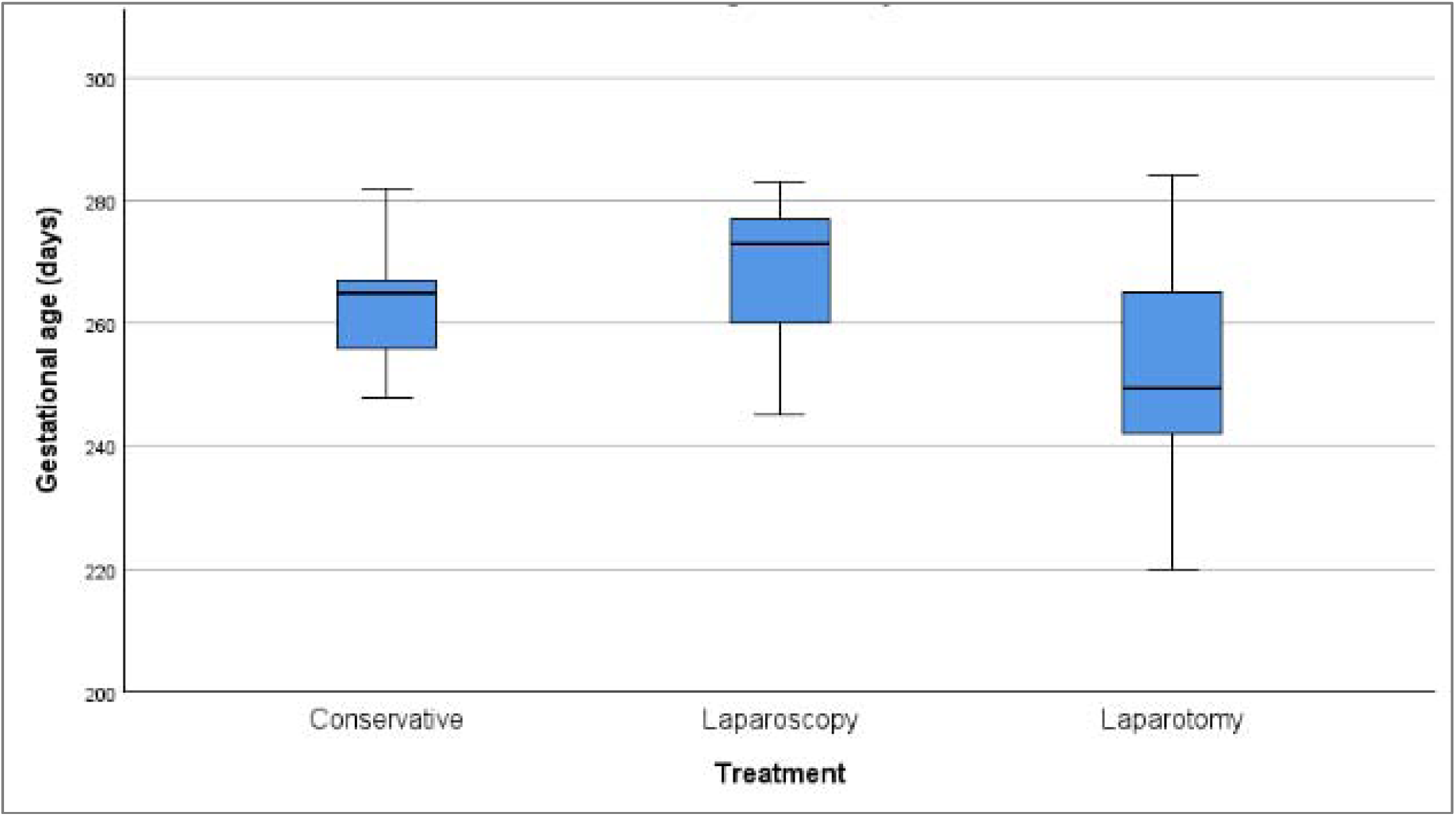
Boxplot of the gestational age at delivery according to complication treatment.

The details of labor and delivery are shown in Supplementary Table 1. Twelve women gave birth during admission for a BS complication, while 21 women were discharged and gave birth during a later admission.

#### Delivery during the same admission (N = 12)

Nine out of 12 births were preterm. The mean gestational age at birth was 35 weeks and 5 days (250 days) (range 220-276). Eight of these underwent a primary c-section (CS) for various indications: four had severe maternal pain requiring urgent laparotomy. The preference for laparotomy over laparoscopy was not specified. Two women underwent a laparoscopic assessment which was converted into a laparotomy due to an unresolved obstruction. This necessitated a simultaneous CS due to the lack of perioperative fetal monitoring and concerns about potential fetal distress. Two additional women had CS prior to further surgical evaluation; one required an urgent category 1 CS due to fetal distress and was later diagnosed with a suture abscess. The other woman experienced increasing discomfort after her scheduled repeat CS, which led to a laparoscopic evaluation the following day.

Four women were hospitalized for observation. Two successful vaginal deliveries were achieved following labor induction, while two secondary CS were performed: one due to failed induction with subsequent laparoscopic gastric band removal, and the other due to intrapartum fetal distress, revealing an IH during CS.

#### Delivery during a later admission (N = 21) (Supplementary Table 1)

The mean gestational age was 38 weeks and 3 days (269 days) (range 247-284 days). There were four preterm births. Most women went into spontaneous labor and gave birth vaginally (n = 9). The reasons for a primary CS (N = 4) were mostly obstetric, except for one case in which it was deemed better to avoid Valsalva maneuvers four weeks after laparotomy.

The CS rate was high (48.5%) and even higher (83.3%) if the delivery took place during the patient’s initial admission. The likelihood of undergoing a CS was 12.5 times higher if delivery occurred during the same admission as the surgical complication, compared to delivery during a later admission (OR = 12.5, 95% CI 2.089 to 74.808). The likelihood of having a vaginal delivery (compared to a CS) increased by 10% for each additional day of gestational age (OR = 1.1, 95% CI 1.027 to 1.178).

### Perinatal outcomes

Birth weight (median 2920g and p30) was within the normal range for 21 newborns. Eight neonates were small for gestational age (SGA, <10^th^ percentile), including three delivered during the same admission and five during a subsequent admission, while four were large for gestational age. Eleven neonates were admitted to the NICU. Reasons for admission were prematurity (N = 7), fetal asphyxia (N = 2), dysmaturity (N = 1), and infection complicated by a pneumothorax (N = 1). The Apgar score was available for 29 children. All had normal 5 min Apgar score, with exception of the two newborns with asphyxia (5 min Apgar score of 5 and 6).

One case of neonatal mortality occurred following an emergency CS for fetal distress. The newborn experienced severe asphyxia at birth (blood pH < 7) and was diagnosed with hypoxic-ischemic encephalopathy. Despite the initiation of therapeutic hypothermia, the death of the newborn was unavoidable.

## Discussion

### Principal findings

This nationwide population-based surveillance study provides data on surgical complications during pregnancy following bariatric surgery in Belgium.

Although uncommon, these complications were associated with substantial obstetric and neonatal morbidity, particularly preterm birth (nearly 40%) and high CS rates (nearly 49%).

Most complications were related to internal herniation after bariatric procedures involving intestinal rerouting and occurred in the third trimester. This timing is consistent with previous reports and may reflect the combined effects of anatomical changes following bariatric surgery and the physiological increase in intra-abdominal pressure during advanced pregnancy.^18^ Although no maternal deaths occurred, bowel resection was required in two women with small bowel strangulation and segmental bowel necrosis.^18^

The mean total weight loss at conception was 36.8%, which is well above the threshold of 20% recommended by the International Federation for the Surgery of Obesity (IFSO).^19^

### Strengths and weaknesses

The strengths of this study include its nationwide scope, prospective case ascertainment through a structured obstetric surveillance system (B.OSS), and the use of national health insurance data to contextualise the observed cases. Nevertheless, underreporting remains a limitation (due to the voluntary nature of reporting and regional discrepancies in participation^20^) and the small number of cases limits statistical power and precludes formal comparisons between bariatric procedures. As such, the findings should be interpreted as descriptive and hypothesis-generating.

### Comparison with existing literature

The observed rate of surgical complications in this study (0.56% overall and 0.88% following RYGB) was lower than previously reported estimates, ranging from 2% to 11% for small bowel obstruction in pregnancies following RYGB documented in small-scale observational studies or case reports.^7, 21, 22^ However, this lower incidence should be interpreted with caution and is most probably underestimated due to underreporting and case ascertainment limitations inherent to surveillance-based studies. Although bariatric surgery remains widely used in women of reproductive age, the growing role of pharmacological weight-loss therapies may modify future patterns of complications in pregnancy after weight loss interventions.^23^

### Clinical implications

A systematic review by Petrucciani et al. reported maternal and perinatal mortality rates of 2,5% and 7,5%, respectively. Delay in diagnosis and intervention were identified as the main contributors to adverse outcomes.^9^ Our findings confirm that clinical symptoms are often non-specific and may overlap with common pregnancy-related complaints, contributing to diagnostic delay. Clinicians must be vigilant for the classical triad of symptoms - abdominal pain, nausea, and vomiting- and avoid misattributing them to normal pregnancy processes, especially in the third trimester. Following RYGB the most dangerous complication is mesenteric strangulation with bowel ischemia, which may manifest as severe abdominal or back pain in the absence of classical obstructive signs. This atypical presentation may further delay diagnosis during pregnancy, with a possible evolution towards septic shock (SIRS).

In our series, MRI suggested the diagnosis in most cases in which it was performed. This finding supports the use of MRI in cases of strong clinical suspicion but should not be interpreted as evidence of superiority over other imaging modalities. Despite previous literature indicating lower sensitivities, MRI is considered equivalent to CT while avoiding fetal radiation exposure^24, 25^.^24, 25^ Nonetheless, given the limited availability and the duration of the examination, resorting to CT is indicated when prompt diagnosis is needed and MRI is unavailable.^23^ Abdominopelvic CT scans in pregnant women deliver an average radiation dose of 24 mGy, which is considered safe as doses below 50 mGy pose a negligible risk to the fetus.^26^

Importantly, inflammatory parameters may be normal or only mildly elevated, which should not reassure clinicians in the presence of persistent pain.^9, 27^ Increased lactate levels can raise suspicion for ischemia and influence the decision toward urgent surgical intervention.^28, 29^ The threshold for a diagnostic laparoscopy should be low, even if imaging is negative but pain is persistent.

Early surgery, ideally within 48 hours of symptom onset, is critical to prevent bowel ischemia.^7^ Given the increased risk of preterm birth in this population, the benefits of corticosteroids for fetal lung maturation should be cautiously balanced against the risks of delaying surgery, ensuring maternal safety remains a priority.

The decision to use a laparoscopic approach is contingent upon the surgeon’s expertise with pregnant patients. Reasons to convert to open surgery include unsuccessful reduction of the obstruction site, extensive bowel resection, or perioperative complications (e.g. intestinal perforation). Additionally, the presence of fetal distress may favor the choice for a laparotomy, allowing for a simultaneous CS. These findings are consistent with recent evidence suggesting that a laparoscopic approach is feasible and often preferable in pregnant women with suspected small bowel obstruction after RYGB. In a recent single-centre series of 32 cases, laparoscopy was associated with favourable maternal outcomes and should be considered the preferred initial approach when surgical expertise is available.^30^ In our cohort, laparoscopy was performed in the majority of cases and remained feasible even in advanced gestation, further supporting this approach.

The interval between bariatric surgery and conception was less than one year in several women. For clinical guideline development groups, the findings emphasize the need for standardized guidelines on the timing of conception. Ideally, conception should be delayed for 12-18 months post-surgery, with close weight loss monitoring. Contraceptive counselling should be incorporated into preoperative consultations, particularly considering the reduced efficacy of oral contraceptives after RYGB,^31^ which makes long-acting reversible contraception highly recommended.^6^ Centralized care, including an obstetric high risk care unit and NICU, can mitigate perinatal morbidity and mortality.^32^

Procedure selection may be relevant for women with a current or future pregnancy wish. While causal inferences cannot be drawn from this study, the absence of observed complications following sleeve gastrectomy and the predominance of complications after bypass procedures underscore the need for balanced counselling. While RYGB has metabolic benefits (resolving dyslipidemia, type 2 diabetes and gastroesophageal reflux disease), there may be a higher pregnancy-related surgical risk.^19, 33, 34^

### Research implications

Further multicenter, population-based studies, ideally coordinated via INOSS, are required to validate these findings internationally and to better define the optimal bariatric procedure for women planning pregnancy.

## Conclusion

Surgical complications during pregnancy in women with a history of bariatric surgery are uncommon but clinically significant. They predominantly occur after bariatric procedures involving intestinal rerouting and are associated with considerable obstetric morbidity despite maternal outcomes. Heightened clinical vigilance, prompt diagnostic evaluation, and timely surgical management remain essential to optimise outcomes for both mother and child.

As increasing numbers of women of reproductive age undergo bariatric surgery, obstetric care providers should maintain a high index of suspicion for surgical pathology when evaluating abdominal complaints during pregnancy.

## Author contribution statement

Paulien De Mulder: validation (supporting), formal analysis (lead), investigation (lead), writing – original draft (lead), writing – review and editing (equal).

Karolien Benoit: methodology (supporting), software (lead), investigation (lead), data curation (lead), writing – review and editing (equal), project administration (lead).

Caroline Daelemans: conceptualization (supporting), methodology (supporting), writing – review and editing (equal), supervision (supporting).

Frédéric Debiève: conceptualization (supporting), methodology (supporting), writing – review and editing (supporting).

Roland Devlieger: conceptualization (supporting), methodology (supporting), writing – review and editing (equal).

Kristien Roelens: conceptualization (supporting), methodology (supporting), writing – review and editing (equal), supervision (supporting), project administration (supporting).

Yves Van Nieuwenhove: writing – review and editing (equal).

Griet Vandenberghe: conceptualization (lead), methodology (lead), validation (lead), formal analysis (supporting), investigation (supporting), writing – original draft (supporting), writing – review and editing (lead), supervision (lead), project administration (lead).

## Patient and public involvement statement

patients and/or the public were not involved in the design, conduct, reporting, or dissemination plans of this research.

## Data availability statement

The data underlying this study are not publicly available due to legal and ethical restrictions. Data are collected within the framework of a national surveillance system and cannot be shared without appropriate data transfer agreements (DTA) and in accordance with the conditions defined in the informed consent. Data may be made available from the authors upon reasonable request and subject to approval by the relevant governance bodies.

## Provenance and peer review

Not commissioned; externally peer reviewed.

## Acknowledgements

The authors would like to thank all local B.OSS investigators for their monthly case reporting, as well as all contributors involved in completing the data collection forms (DCF). All members of the B.OSS Collaborating Group contributed to data collection and monthly case reporting, and meet the criteria for group authorship.

## Appendix 1

Members of the B.OSS Collaborating Group

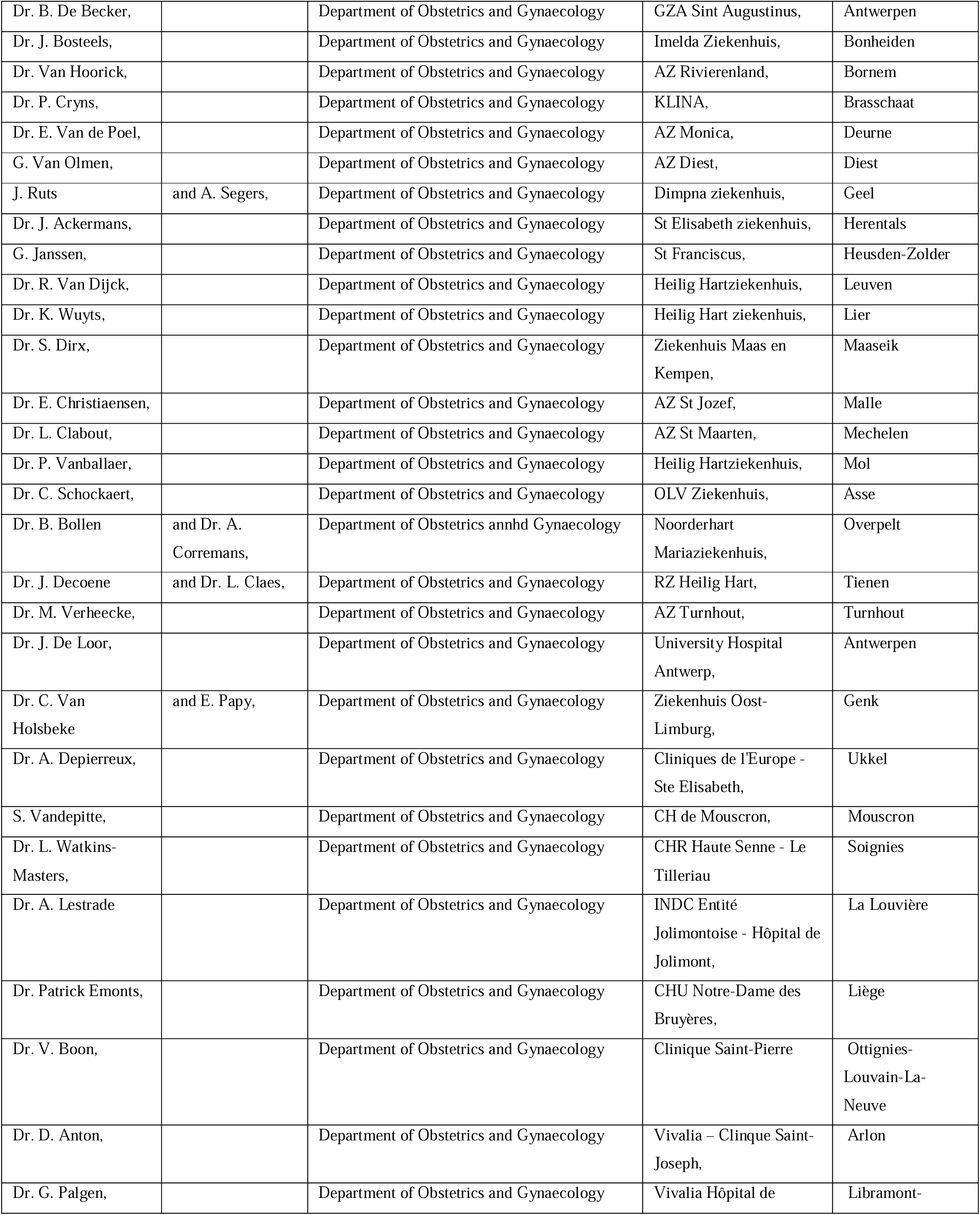
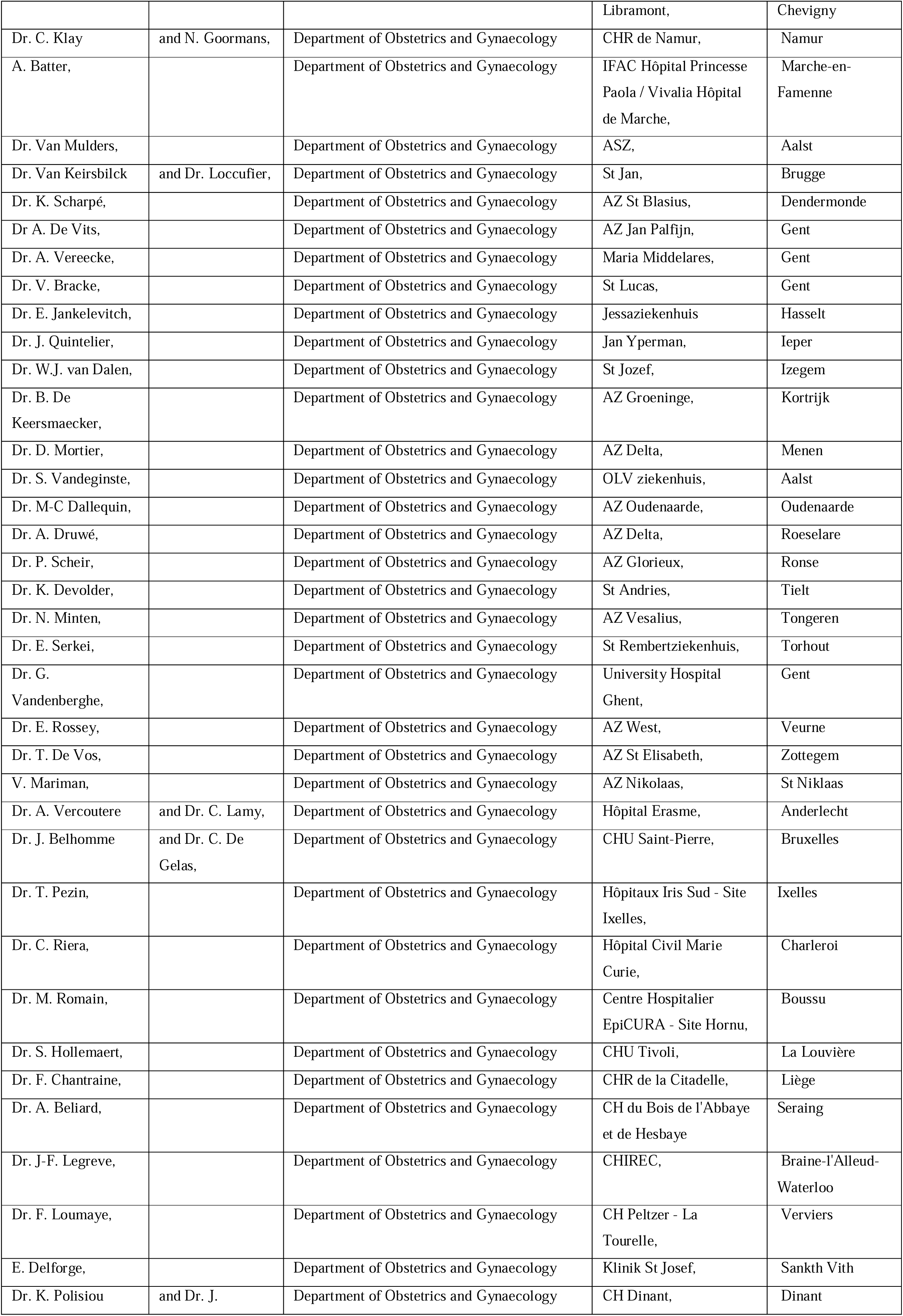
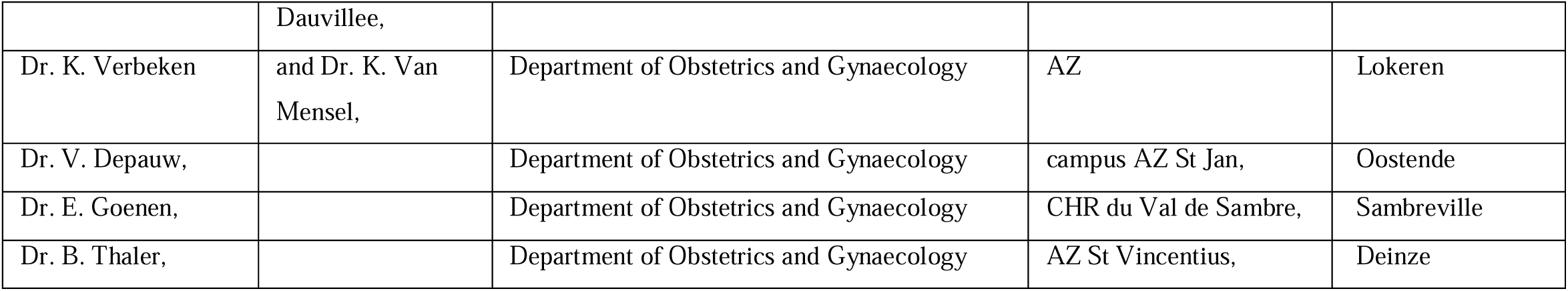

---

**Supplementary Table 1:**
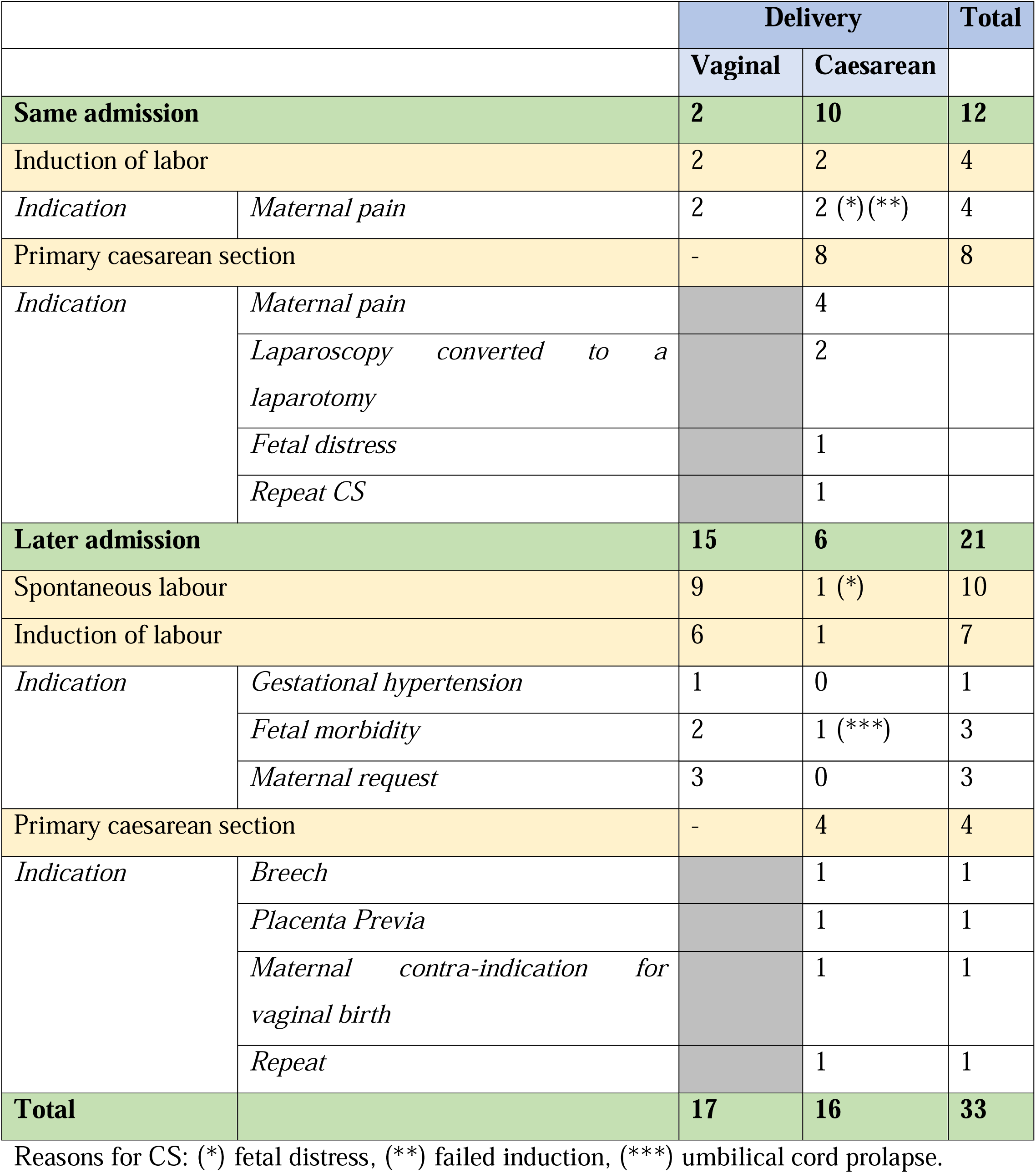
Labor and delivery in women with surgical complications of BS in pregnancy.

|  |  | Delivery |  | Total |
| --- | --- | --- | --- | --- |
|  |  | Vaginal | Caesarean |  |
| **Same admission** |  | **2** | **10** | **12** |
| Induction of labor |  | 2 | 2 | 4 |
| *Indication* | *Maternal pain* | 2 | 2 (*)(**) | 4 |
| Primary caesarean section |  | - | 8 | 8 |
| *Indication* | *Maternal pain* |  | 4 |  |
|  | *Laparoscopy converted to a laparotomy* |  | 2 |  |
|  | *Fetal distress* |  | 1 |  |
|  | *Repeat CS* |  | 1 |  |
| **Later admission** |  | **15** | **6** | **21** |
| Spontaneous labour |  | 9 | 1 (*) | 10 |
| Induction of labour |  | 6 | 1 | 7 |
| *Indication* | *Gestational hypertension* | 1 | 0 | 1 |
|  | *Fetal morbidity* | 2 | 1 (***) | 3 |
|  | *Maternal request* | 3 | 0 | 3 |
| Primary caesarean section |  | - | 4 | 4 |
| *Indication* | *Breech* |  | 1 | 1 |
|  | *Placenta Previa* |  | 1 | 1 |
|  | *Maternal contra-indication for vaginal birth* |  | 1 | 1 |
|  | *Repeat* |  | 1 | 1 |
| **Total** |  | **17** | **16** | **33** |

Reasons for CS: (\*) fetal distress, (\*\*) failed induction, (\*\*\*) umbilical cord prolapse.

**Supplementary Table 2:**
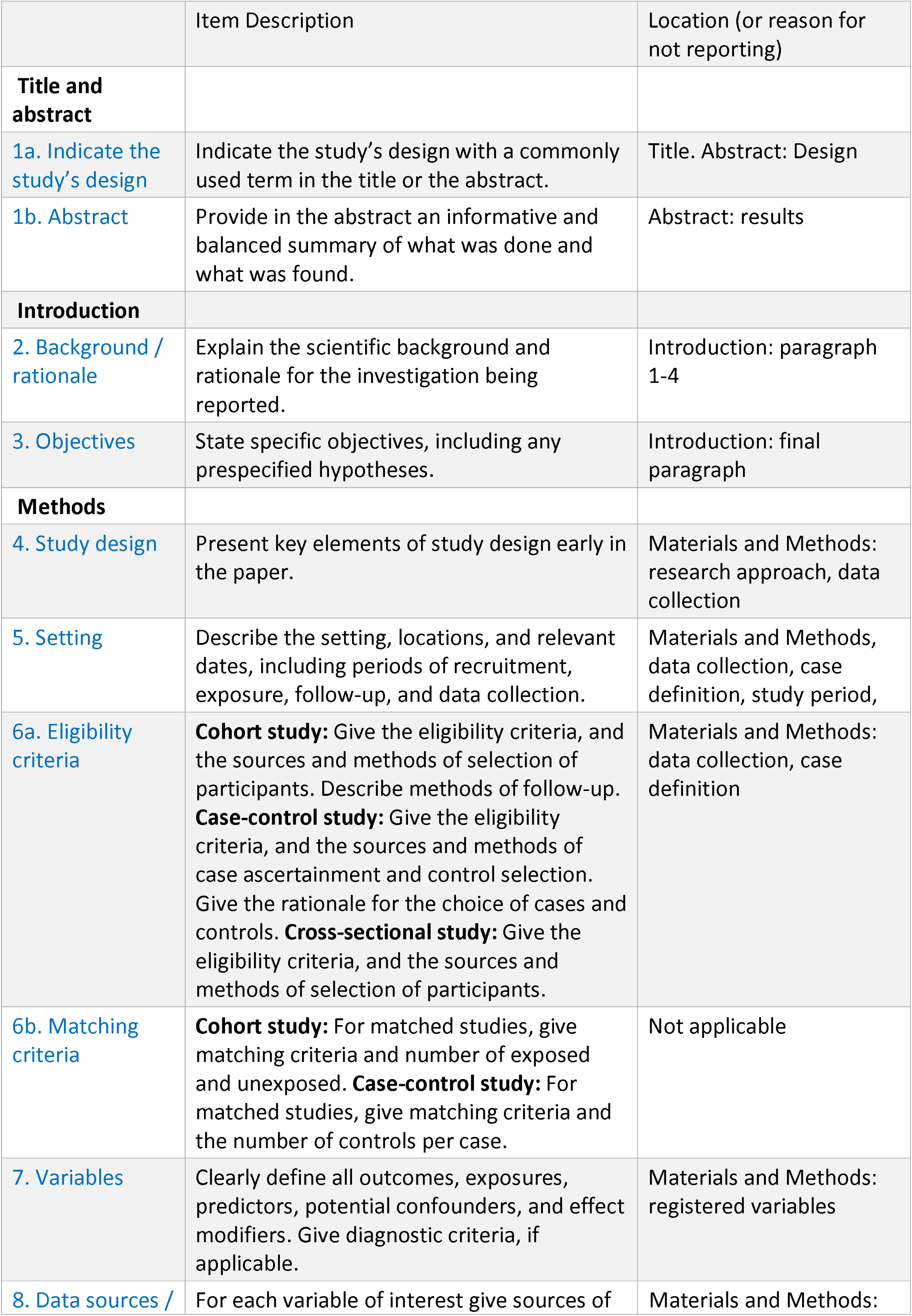
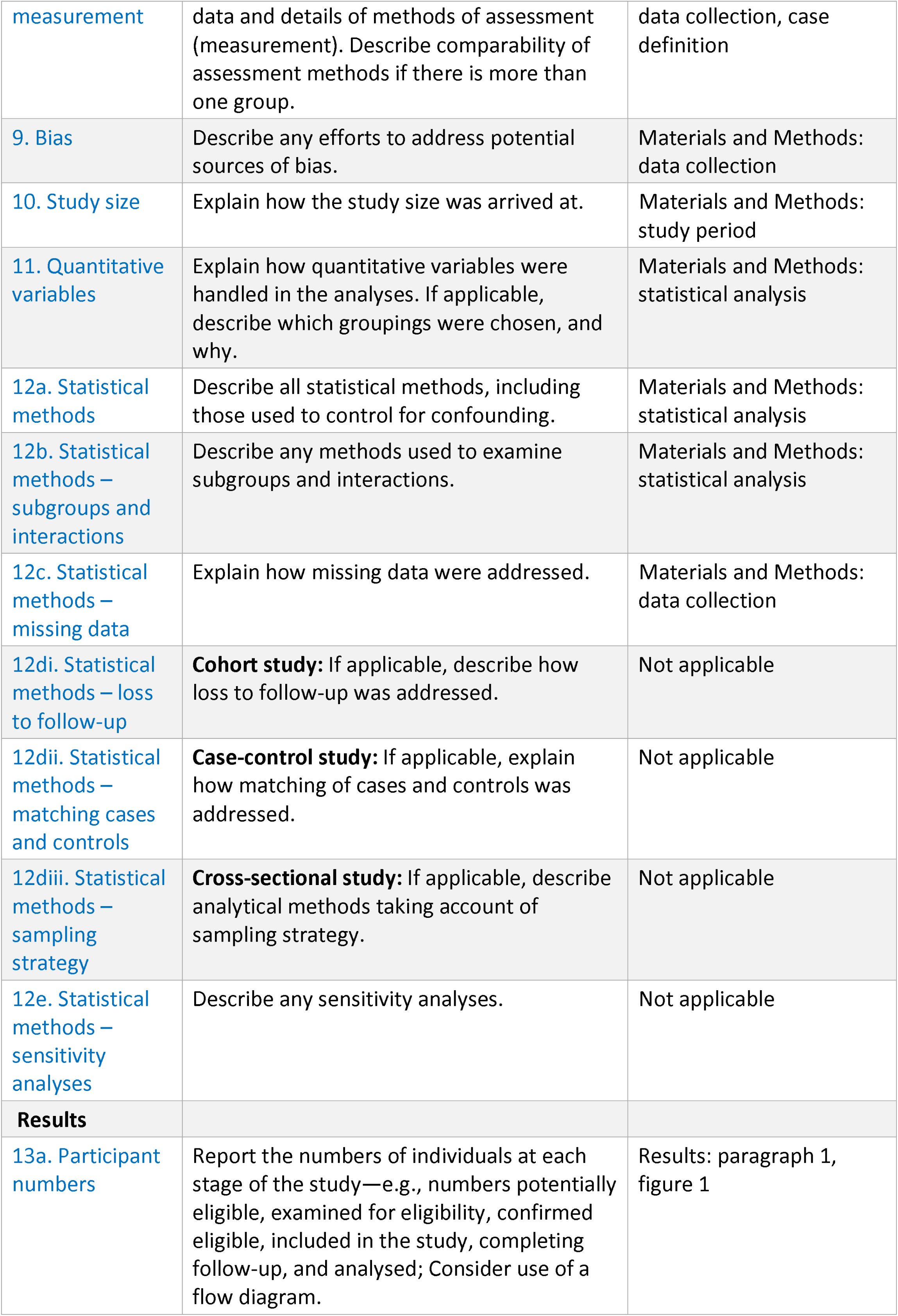
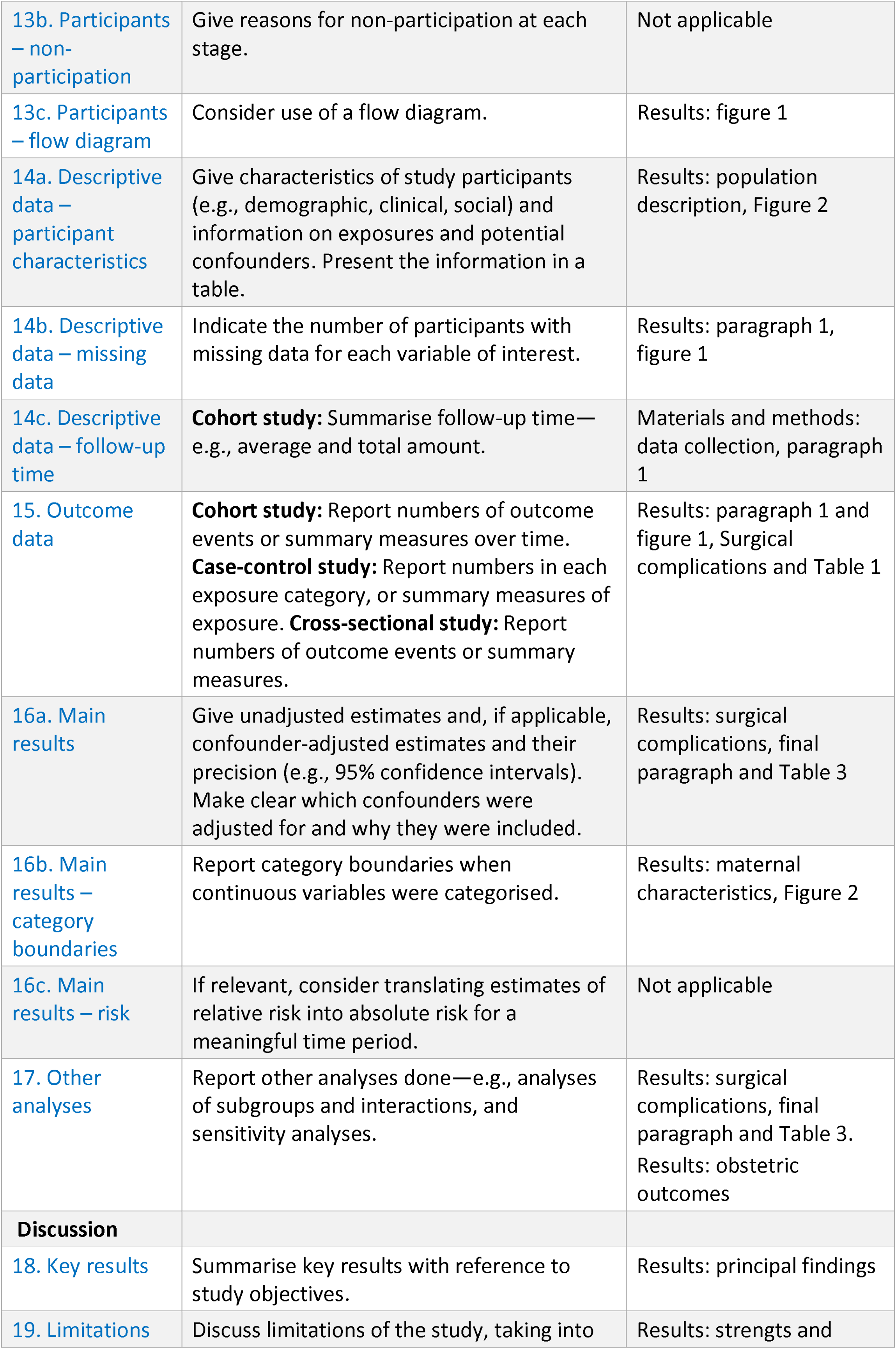
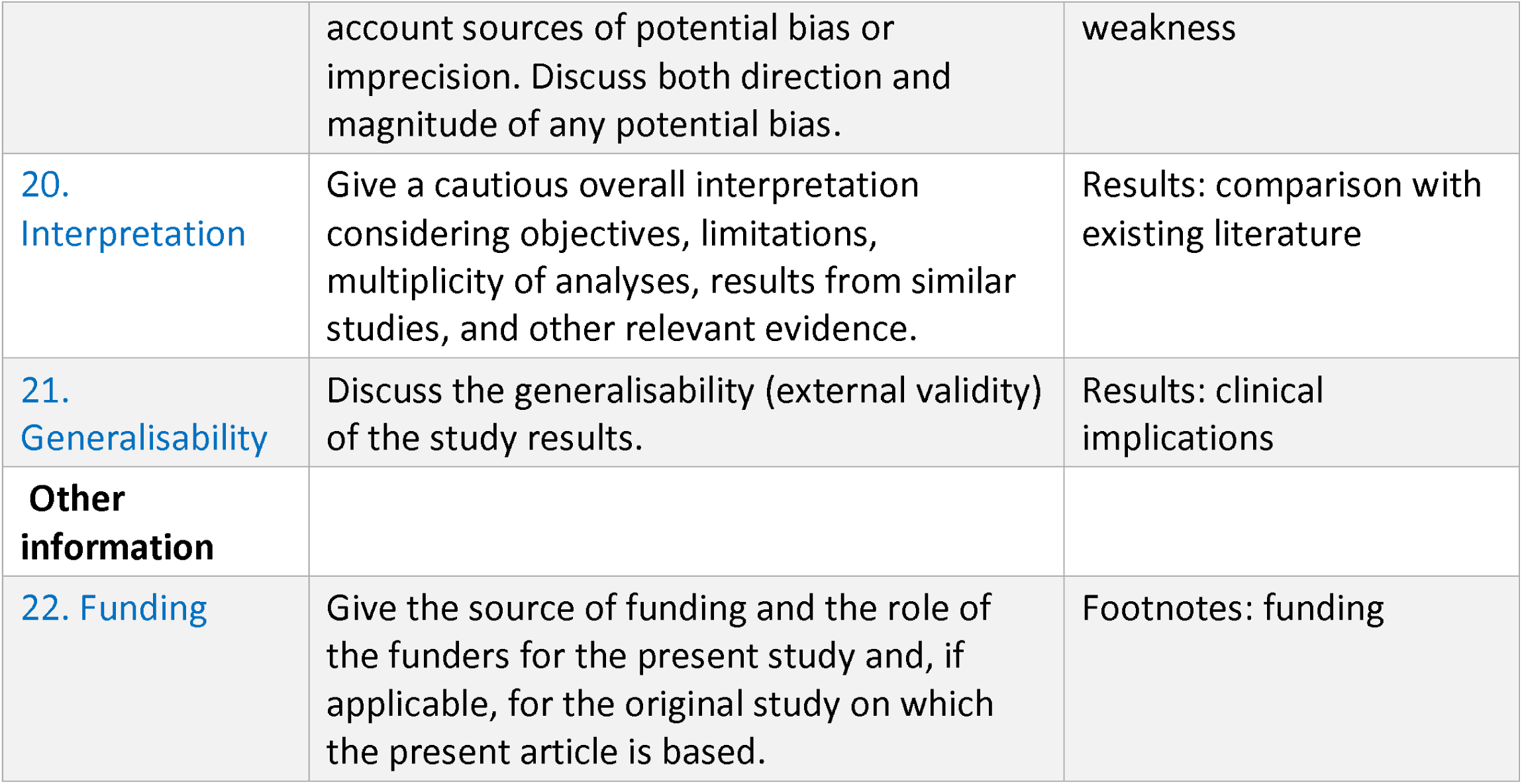
The STROBE reporting checklist.

